# Associations between multiple long-term conditions and mortality in diverse ethnic groups

**DOI:** 10.1101/2022.01.13.22268828

**Authors:** Mai Stafford, Hannah Knight, Jay Hughes, Anne Alarilla, Luke Mondor, Anna Pefoyo Kone, Walter Wodchis, Sarah R Deeny

## Abstract

**Background:** Multiple conditions are more prevalent in some minoritised ethnic groups and are associated with higher mortality rate but studies examining differential mortality once conditions are established is US-based. Our study tested whether the association between multiple conditions and mortality varies across ethnic groups in England.

**Methods and Findings:** A random sample of primary care patients from Clinical Practice Research Datalink (CPRD) was followed from 1^st^ January 2015 until 31^st^ December 2019. Ethnicity, usually self-ascribed, was obtained from primary care records if present or from hospital records. Cox regression models were used to estimate mortality by number of long-term conditions, ethnicity and their interaction, with adjustment for age and sex for 532,059 patients with complete data.

During five years of follow-up, 5.9% of patients died. Each additional long-term condition at baseline was associated with increased mortality. This association differed across ethnic groups. Compared with 50-year-olds of white ethnicity with no conditions, the mortality rate was higher for white 50-year-olds with two conditions (HR 1.77) or four conditions (HR 3.13). Corresponding figures were higher for 50-year-olds of Black Caribbean ethnicity with two conditions (HR=2.22) or four conditions (HR 4.54). The direction of the interaction of number of conditions with ethnicity showed higher mortality associated with long-term conditions in nine out of ten minoritised ethnic groups, attaining statistical significance in four (Pakistani, Black African, Black Caribbean and Black other ethnic groups).

**Conclusions:** The raised mortality rate associated with having multiple conditions is greater in minoritised ethnic groups compared with white people. Research is now needed to identify factors that contribute to these inequalities. Within the health care setting, there may be opportunities to target clinical and self-management support for people with multiple conditions from minoritised ethnic groups.

## Introduction

The number of people with multiple long-term conditions, or multimorbidity, is rising. Though there is no accepted international definition of multimorbidity(1), recent large-scale studies using electronic health records in the UK estimate 23-27% of people have two or more long-term conditions(2)(3). Multimorbidity has been consistently associated with poorer outcomes for patients, with risk of death increasing with each additional condition(4)(5) and some studies suggesting that the association may be multiplicative(6), and more pronounced when conditions concern different body systems (complex multimorbidity)(7)(8)(9)(10)(11) though the association between higher risk of mortality and multimorbidity is weaker at older ages (12)(13). Multimorbidity is also associated with higher use of health care(2) and reduced quality of life(14,15). Many health and care systems are designed to care for patients with single conditions, but there is growing recognition that if they are to improve outcomes for patients, health care systems must be adapted to address the challenge of multimorbidity(16)(17).

There is an established body of evidence that the prevalence of multimorbidity is socially patterned. Previous studies have demonstrated an association between the prevalence of multiple conditions and socioeconomic disadvantage in households (18) and local areas (2,3)(19). The prevalence of multiple conditions is also higher in some minoritised ethnic groups(20). People from some minoritised ethnic groups are more likely to have experienced discrimination and multiple disadvantage over their life course, leading to an increased risk of experiencing material deprivation, living in deprived areas and an associated higher prevalence of downstream behavioural risk factors including smoking and obesity(20)(21). Poorer experience of healthcare services has also been reported by some minoritised ethnic groups, and they are less likely than their counterparts in the majority population to report that they are able to manage their own health(22).

Whether minoritised ethnic groups experience disadvantage or discrimination over the course of their lives will differ between countries and over time, and as a result, evidence on mortality risk for minoritised ethnic groups varies. For example while excess deaths in Black populations in the US have remained high for many years (23), in the UK, the link between ethnic minority status and mortality risk varies by cause of death (24) and migration status (25). Research in the UK on this topic has been hampered due to lack of ethnicity data on death certificates and historically poor recording of ethnicity in medical records, though the latter has improved markedly in recent years.

A previous study in the United States of America found that having multiple chronic conditions resulted in reduced life expectancy but the impact did not differ between African Americans and non-Hispanic white people(26). Analysis of the Health and Retirement Study, on the other hand, found that Black and Hispanic Americans were more likely to have multisystem multimorbidity and more likely to die during follow-up compared with their White American counterparts(27). We are not aware that this has been assessed in the UK context, though here studies have investigated ethnic differences in long-term survival for people with a single or index condition of interest. These point to the possibility of ethnic differences in survival across a range of conditions, for example lower two-year survival for Black women in England with breast cancer(28) and higher survival for people in London with unipolar depression from Black Caribbean, Black African, South Asian and Chinese ethnic backgrounds(29).

It is plausible that many of the factors contributing to high prevalence of multiple conditions may also contribute to poorer survival in minoritised ethnic groups once multiple conditions are established. Likewise, the potential for differences across ethnic groups in the association between long-term conditions and mortality may be greater where the organisation of health care and the recommended treatments and lifestyle changes are especially complex, as is the case with multiple conditions.

The aims of our study were to estimate the mortality risk of having multiple conditions, assess whether this risk is seen for complex multimorbidity, and examine whether the magnitude or direction of these risks varies across ethnic groups, compared with people of white ethnicity living in England.

## Methods

### Participants

Our sample was drawn from primary care records. Over 95% of the England population are registered with a general practice. A random sample of 600,000 adults (age 18 and over) was drawn from the Clinical Practice Research Datalink (CPRD Aurum (30)). This research database of anonymised routinely collected primary care records captures diagnoses, symptoms, prescriptions, referrals and tests and includes over 40 million patients (13 million currently registered as of June 2021). CPRD Aurum comprises GP practices using the EMIS Web software (one of four main general practice IT systems in operation) that have agreed to contribute data. Eligible adults were in a CPRD practice on 1^st^ January 2014 (to ensure records were up to date at least one year before the study start), were alive and still registered at the study start on 1^st^ January 2015, and were eligible for linkage to Hospital Episode Statistics (HES) and Office for National Statistics mortality data. They were followed until the study end (31^st^ December 2019) or death if this was earlier and were censored if they left the CPRD practice or the practice stopped providing data to CPRD. The study was approved by the CPRD team (eRAP protocol number 20_000239).

### Measures

Survival time was calculated from 1^st^ January 2015 to death or censoring.

The number of long-term conditions was counted at study start. We used a list of 32 physical and mental health conditions (Supplementary Table 1) that have previously been associated with higher mortality risk, poorer functioning, and requiring primary care input (2,3). This was repeated to calculate number of conditions at the study end or censoring date, which may be fewer than at study start as we allowed for three conditions (anxiety/depression, asthma and cancers) to resolve.

Complex multimorbidity was defined as having three or more long-term conditions in three or more different body systems(31) (Supplementary Table 1).

Ethnic identity, usually self-ascribed, was obtained from SNOMED codes recorded by the GP or, where that was missing or incomplete (29.8%), from linked HES (Hospital Episode Statistics) records. Where multiple values of ethnicity have been recorded, we selected the modal value where this was unique, or the most recent value(32). Categories from the England 2011 census were used in our analysis but we combined white British, white Irish and other white because these separate categories were not available in HES.

Socioeconomic deprivation was captured by 2015 Index of Multiple Deprivation (IMD) decile in the patient’s area of residence based on lower-level super output area boundaries.

### Statistical analysis

The analytical sample included those with complete data on sex, age (n=0 excluded), ethnicity (n= 67524 excluded), or deprivation (n=417 excluded). Excluded patients were younger, more likely to be men, over-represented in less deprived areas, and had fewer conditions (Supplementary Table 2).

The association between survival time and baseline number of long-term conditions was modelled using a multilevel Cox proportion hazards model with adjustment for baseline age (centred at age 50 to aid interpretation), sex and ethnicity. Number of conditions was included as a continuous variable after confirming its association with survival time was linear (Supplementary Table 3). Ethnicity by age interactions were included as this improved model fit. A two-level model was used to allow for the clustering of patients within GP practices (model 1).

We assessed model 1 for violations of the proportional hazards assumption. The association between sex and mortality hazard was found to depend on follow-up time (p=0.004), with a marginally higher hazard for men after 2.5 years of follow-up. The association between baseline age and mortality hazard also depended on follow-up time (p=0.02), with a marginally higher hazard with advancing age after 2.5 years of follow-up. However, the differences across follow-up time were small. Furthermore, allowing for time-varying estimates for sex and age did not alter the estimates for the main variables of interest (namely, ethnicity and number of long-term conditions) so we elected to present the simpler model without time-varying estimates.

To examine whether the association between survival time and long-term conditions varied by ethnicity, we added ethnicity by number of conditions interaction terms (model 2). A likelihood ratio test was used to test the combined statistical significance of these interactions (model 2 vs model 1).

We examined two possible factors that could explain survival differences across ethnic groups, if any were observed (model 3). We added number of long-term conditions at end of follow-up. Patterns and rate of long-term condition acquisition vary across ethnic groups (20,27,33)(34). We also added socioeconomic deprivation. There is a well-established relationship between ethnic minority identity and greater socioeconomic deprivation, driven by long-standing structural factors that disadvantage people from minoritised ethnic groups in multiple domains including housing, education and employment. We hypothesised that any ethnic differences in the association between survival time and baseline number of conditions would be smaller in models that included number of long-term conditions at end of follow-up and deprivation.

In sensitivity analysis, we repeated model 2 replacing baseline number of conditions with presence or absence of complex multimorbidity. We also looked at the presence of conditions in specific body systems that are leading causes of death (35) and that had sufficient sample size across ethnic groups (endocrine, circulatory, respiratory). Here, significance refers to statistical significance at the 5% level.

## Results

During the 5-year follow-up period, 5.9% of patients died (Table 1). The majority of patients were of white ethnic background (85.4%) and these were older and over-represented in less deprived areas compared with all other ethnic groups (Supplementary Table 4). The unadjusted mean number of long-term conditions at baseline was highest in the white ethnic group (1.23) and lowest in the Chinese ethnic group (0.33).

**Table 1.**
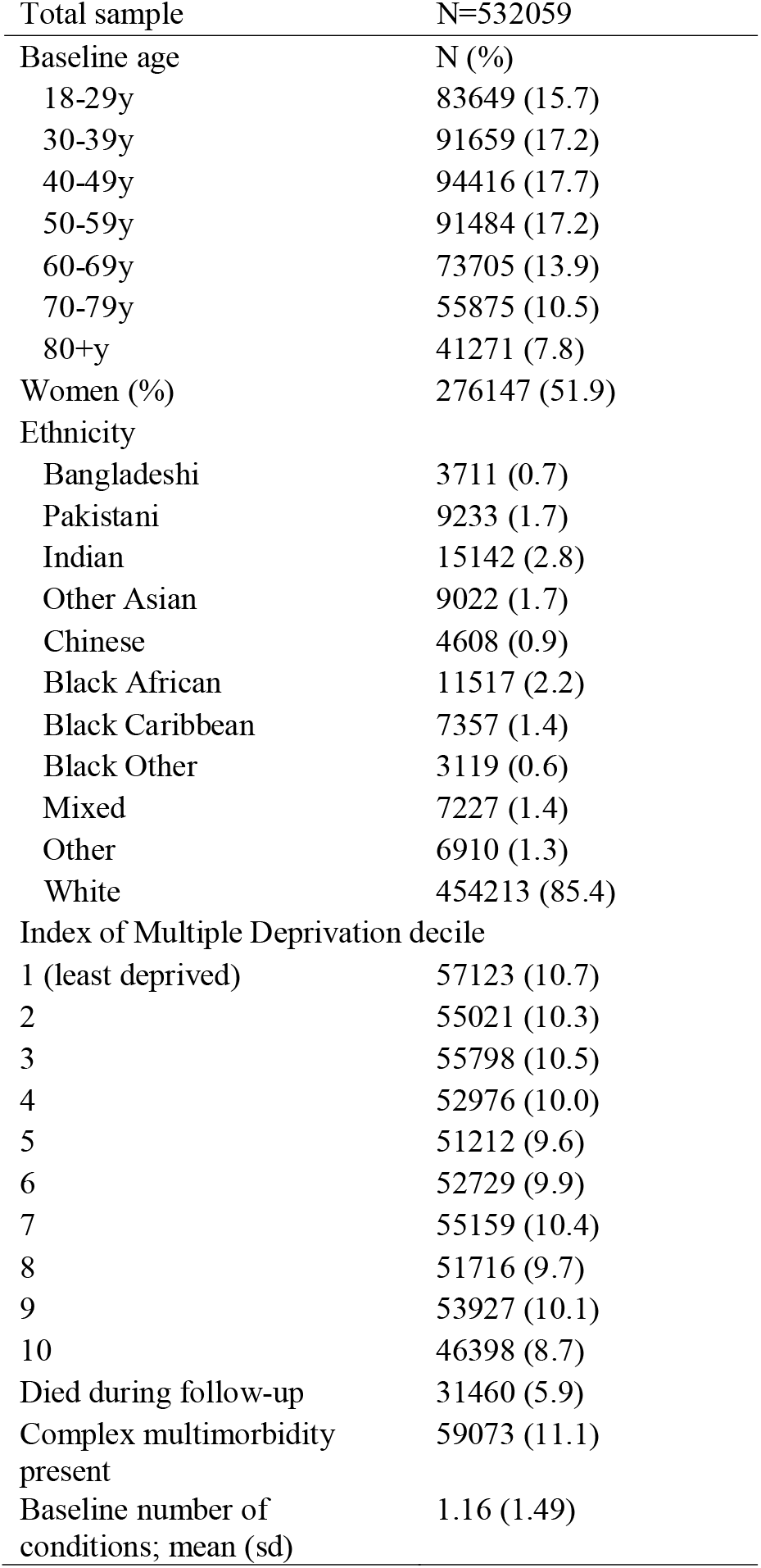
Description of the analytical sample.

The initial model (model 1) addressed the first objective, to assess whether number of long-term conditions is associated with mortality. This model is based on the assumption that the association between number of conditions and mortality is consistent across ethnic groups. Each additional long-term condition at baseline was associated with increased mortality. For example, the hazard ratio (HR) was 1.80 with two conditions and HR=3.25 with four conditions, compared to the reference group with no conditions at age 50 (example hazard ratios in Table 2 and Supplementary Table 5 for full set of estimates). Statistically significant interactions for number of conditions by baseline age shows that the relative difference in mortality for those with more versus no conditions was smaller at older ages.

**Table 2.** Mortality hazard ratios at specific ages by baseline number of conditions or by ethnicity (assuming association between number of conditions and mortality is same for all ethnic groups)^a^.

|  | Age 50 | Age 60 | Age 70 |
| --- | --- | --- | --- |
| Baseline number of conditions |  |  |  |
| 0 | **1** | 2.73 | 7.44 |
| 1 | 1.34 | 3.55 | 9.37 |
| 2 | 1.80 | 4.61 | 11.79 |
| 3 | 2.42 | 5.99 | 14.84 |
| 4 | 3.25 | 7.79 | 18.67 |
| Ethnicity |  |  |  |
| Bangladeshi | 0.86 | 2.24 | 5.88 |
| Pakistani | 1.08 | 2.68 | 6.65 |
| Indian | 0.62 | 1.81 | 5.30 |
| Other Asian | 0.74 | 1.93 | 5.00 |
| Chinese | 0.40 | 1.34 | 4.41 |
| Black African | 0.97 | 2.26 | 5.30 |
| Black Caribbean | 1.21 | 2.87 | 6.83 |
| Black other | 1.31 | 2.64 | 5.34 |
| Mixed | 1.22 | 2.62 | 5.64 |
| Other | 0.78 | 2.11 | 5.67 |
| White | **1** | 2.73 | 7.44 |

<sup>a</sup>Estimates are from Cox regression model 1 including baseline number of conditions, ethnicity, baseline age, sex, and interactions for number of conditions by age and ethnicity by age

Ethnicity was associated with mortality and this association depended on age. Taking patients of white ethnicity age 50 years at baseline as the reference, the hazard ratio was above 1 indicating a higher relative mortality rate for the Pakistani, Black Caribbean, Black other and mixed ethnic groups. For 70-year olds, the hazard ratio was highest for those of white ethnicity. At all ages, the mortality rate was significantly lower for those of Indian or Chinese ethnicity than for those of white ethnicity.

Model 2 summarises analysis addressing the second objective and provides evidence that the association between number of conditions and mortality differed across ethnic groups (likelihood ratio test 0.05<p<0.01; example hazard ratios in Table 3). Compared with 50-year-olds of white ethnicity with no conditions, the mortality rate was higher for white 50-year-olds with two conditions (HR 1.77) or four conditions (HR 3.13). The corresponding figures were higher for 50-year olds of Black Caribbean ethnicity with two conditions (HR=2.22) or four conditions (HR 4.54). Positive estimates for the interaction of number of conditions with ethnicity show that the higher mortality associated with having more conditions was greater in all minoritised ethnic groups, with the exception of those from a Bangladeshi background (full set of estimates in Supplementary Table 6). Interaction terms attained statistical significance for Pakistani, Black African, Black Caribbean and Black other ethnic groups (Figure 1).

**Table 3.** Mortality hazard ratios at specific ages by baseline number of conditions and ethnicity (examining whether ethnicity moderates the association between total number of LTCs and survival)^a^.

|  | Age 50 |  |  | Age 70 |  |  |
| --- | --- | --- | --- | --- | --- | --- |
|  | 0 conditions | 2 conditions | 4 conditions | 0 conditions | 2 conditions | 4 conditions |
| Ethnicity |  |  |  |  |  |  |
| Bangladeshi | 0.92 | 1.52 | 2.51 | 6.58 | 9.55 | 13.87 |
| Pakistani | 0.82 | 1.87 | 4.26 | 4.29 | 9.79 | 22.35 |
| Indian | 0.58 | 1.12 | 2.15 | 4.74 | 9.13 | 17.62 |
| Other Asian | 0.67 | 1.37 | 2.81 | 4.10 | 8.40 | 17.21 |
| Chinese | 0.37 | 0.78 | 1.61 | 3.76 | 7.81 | 16.23 |
| Black African | 0.84 | 1.84 | 4.05 | 4.12 | 9.06 | 19.91 |
| Black Caribbean | 1.09 | 2.22 | 4.54 | 5.66 | 11.58 | 23.67 |
| Black other | 0.94 | 2.56 | 6.95 | 2.94 | 7.98 | 21.64 |
| Mixed | 1.11 | 2.24 | 4.52 | 4.82 | 9.74 | 19.67 |
| Other | 0.69 | 1.50 | 3.28 | 4.48 | 9.80 | 21.42 |
| White | 1 | 1.77 | 3.13 | 7.52 | 11.68 | 18.15 |

<sup>a</sup>Estimates are from Cox regression model 2 including baseline number of conditions, ethnicity, baseline age, sex, and interactions for number of conditions by age and ethnicity by age and number of conditions by ethnicity

Model 3 explored the contribution of deprivation and number of conditions at end of follow-up as possible explanations for ethnic differences in the association between long-term conditions and mortality. Increasing deprivation was associated with higher mortality (HR 1.74 95% CI (1.66,1.83) in the most compared with the least deprived tenth of areas in England; Supplementary Table 6). The number of conditions present at the end of follow-up varied little across the ethnic groups (Supplementary Table 4) and was not associated with mortality independently of all other covariates (HR 1.00 95% CI (0.99,1.01) for each additional condition). Inclusion of these two variables did not materially alter the estimates for ethnicity by number of baseline conditions interactions. In other words, there was no evidence that area deprivation and onset of new conditions are part of the underlying explanation for ethnic differences in the link between multiple conditions and mortality. We note that there was evidence that the main effect association between ethnicity and mortality was suppressed when deprivation was ignored. Lower mortality rates for most minoritised ethnic groups compared to the white ethnic group were even lower once deprivation was included in the regression model.

Complex multimorbidity was associated with higher mortality risk and the magnitude of this association also varied across ethnic groups and age (Table 4 and Supplementary Table 7). Compared with 50-year-olds of white ethnicity without complex multimorbidity, mortality was higher for those of white ethnicity with complex multimorbidity (HR 2.35) and higher still for people with complex multimorbidity of Pakistani ethnicity (HR 3.48) or Black Other ethnicity (HR 6.66). All ethnicity by complex multimorbidity interaction estimates were positive, with the exception of the mixed ethnic group, indicating a higher mortality rate associated with multimorbidity in minoritised ethnic groups compared with people of white ethnicity. The mortality rate associated with complex multimorbidity was significantly raised for people of Pakistani, Chinese, Black African or Black other ethnicity.

**Table 4.** Mortality hazard ratios at specific ages by complex multimorbidity and ethnicity^a^.

| Ethnicity | Age 50 |  | Age 70 |  |
| --- | --- | --- | --- | --- |
|  | Not complex multimorbidity | Complex multimorbidity | Not complex multimorbidity | Complex multimorbidity |
| Bangladeshi | 0.84 | 1.98 | 6.13 | 11.64 |
| Pakistani | 0.94 | 3.48 | 5.56 | 16.48 |
| Indian | 0.62 | 1.47 | 5.46 | 10.40 |
| Other Asian | 0.67 | 1.98 | 4.63 | 10.91 |
| Chinese | 0.35 | 3.04 | 4.40 | 11.94 |
| Black African | 0.82 | 2.85 | 6.72 | 13.09 |
| Black Caribbean | 1.15 | 6.66 | 2.82 | 13.39 |
| Black other | 0.89 | 1.54 | 3.40 | 16.86 |
| Mixed ethnicity | 1.17 | 2.20 | 6.02 | 9.07 |
| Other ethnicity | 0.69 | 1.99 | 5.10 | 11.78 |
| White | 1 | 2.35 | 7.70 | 12.85 |

<sup>a</sup>Estimates are from Cox regression including complex multimorbidity at baseline, ethnicity, baseline age, sex, and interactions for complex multimorbidity by age and ethnicity by age and complex multimorbidity by ethnicity

The presence of one or more circulatory conditions was associated with higher mortality, and the magnitude of this association was significantly greater in people of Pakistani and Black other backgrounds (Supplementary Table 8). At age 50, the hazard ratio for a circulatory condition was 1.12 in the white ethnic group, 1.70 in the Pakistani ethnic group and 2.16 in the Black other ethnic group, compared with people of white ethnicity without a circulatory condition. The presence of one or more endocrine conditions was associated with higher mortality, and the magnitude of this association was significantly greater in people of Pakistani, Indian, Black African and Black other backgrounds. While 50-year-olds of white ethnicity with an endocrine condition had a mortality rate 1.54 times that of their white counterparts with no endocrine condition, the hazard ratio rose to 3.53 for 50-year-olds of Black other ethnicity with an endocrine condition. The presence of one or more respiratory conditions was also associated with higher mortality, though this association was significantly weaker in those from Black African or Black Caribbean ethnic groups.

## Discussion

### Contribution of our study

Our analysis of data from over half a million patients in primary care confirms that having multiple long-term conditions is associated with higher mortality. This association is seen for each additional condition and for combinations of three or more conditions in three different body systems. The raised mortality rate associated with having multiple conditions is greater in minoritised ethnic groups compared with white people. At age 50, the mortality rate for people of white ethnicity with four conditions was 3.13 times that of their counterparts with no conditions. The mortality rate for people of Black Caribbean ethnicity was 4.54 times that of people of white ethnicity with no conditions. At age 70, the corresponding figures were 18.15 for those of white ethnicity with four conditions and 23.67 for those of Black Caribbean ethnicity with four conditions. Our study identified people with multiple conditions from Black African, Black Caribbean, Black other or Pakistani backgrounds as having poorer survival compared with their white counterparts both for total number of conditions and for complex multimorbidity, though there was some variation in mortality risk across the age groups.

Socioeconomic deprivation is strongly associated with mortality risk. People from most minoritised ethnic groups experienced higher levels of deprivation and omitting deprivation from models suppressed some of their mortality advantage. However, deprivation did not contribute to explaining the ethnic differences in the impact of long-term conditions on mortality. As clinicians and policy makers attempt to address the impact of the rising prevalence of multimorbidity, our findings suggest that there is an urgent need to focus on the needs of patients from minoritised ethnic groups as well as those in socioeconomically deprived areas.

### Possible mechanisms

There are some plausible mechanisms which may explain the associations found in our study. Socioeconomic deprivation shows a clear patterning across ethnic groups and could contribute to poorer survival once multiple conditions are established. The role of socioeconomic deprivation was partially tested in this study, using an area-based measure. It remains a possibility that employment status, financial difficulties and other social and economic stressors may contribute to how effectively people from difference ethnic groups can manage their long-term conditions.

Although the National Health Service provides care that is free at the point of need, differences in access to care and satisfaction with care across ethnic groups have been identified in some parts of the health system(36) as accounts of lived experience testify(37). Poorer access, experience and outcomes of preventive health care interventions have been reported among minoritised ethnic groups, though these vary by setting(36). Furthermore, for specific conditions such as diabetes, studies have demonstrated persistent disparities in appropriate management, treatment and outcomes in primary care (38)(39). However, our study did not set out to test the role of health care quality in any ethnic differences we identified.

Other possible explanations for differences in survival across ethnic groups include differences in the combination of conditions that are present. There is evidence that some combinations are more lethal than others. Complex cardiometabolic multimorbidity has been associated with high mortality risk (11). Analysis of Danish register data showed that pairs including cancer, cardiovascular, lung, mental health or a neurological condition had highest mortality risk (7). On the other hand, it has been hypothesised that multimorbidity including a condition that increases the opportunity for care from a physician may result in comorbid diseases being detected and managed, with a corresponding weaker impact on survival(8).

Investigation of condition clusters was beyond the scope of this study, but our analysis showed that the poorer survival for most minoritised ethnic groups was also seen when we focused only on conditions in the circulatory or endocrine systems. This was not seen for conditions in the respiratory system. The small number of people with cancer in most minoritised ethnic groups meant that we could not examine cancers separately but also suggests that cancers are not principal drivers of the ethnic inequalities observed.

### Comparison with previous research

The two previous studies exploring modification of the association between multiple conditions and mortality by ethnicity provided contrasting evidence. Both were based on cohort studies in the US but differed in their measurement of multiple conditions and analytical approach. Our findings align with the study focused on multisystem multimorbidity (27) but further replication is needed, along with international comparative studies to explore the role of societal context.

## Strengths and limitations

A strength of our study is the large sample size which enabled us to disaggregate into eleven ethnic groups. However, there may be differences within groups, notably the white ethnic group as this combined white British, white Irish and other white people. We were unable to investigate possible disparities for minoritised white ethnic groups. In addition, those from other white backgrounds tend to be more socioeconomically disadvantaged compared to the white British group(40) so our study may underestimate differences between the white British majority and non-white minoritised ethnic groups. Usable ethnicity data was complete for almost 90% of the sample, providing considerable scope for analysis to understand ethnic inequalities in health and care. This is an improvement over the last two decades and initiatives to improve ethnicity data quality should raise this even further. Nevertheless, 11% of the sample was excluded due to missing or withheld ethnicity data. Those without ethnicity data had considerably fewer conditions and a lower percentage died during follow-up so would not contribute large number of events to survival models though we cannot rule out the possibility that the association between multiple conditions and mortality is not the same for those excluded.

We included five years of follow-up and ended the study before the start of the Covid-19 pandemic as our focus was on inequalities in a period of usual care. It is well-documented that Covid-19 mortality risk was highest for people from minoritised ethnic groups and people with existing long-term conditions(41). Our analysis may underestimate ethnic differences in the impact of multiple conditions in the context of Covid-19.

Reverse causality due to diagnosis of conditions when a person is seriously ill is one possible explanation for an association between number of conditions and mortality risk. We are not aware of evidence that this bias differs across ethnic groups but cannot rule out this possibility. Other limitations of our study include lack of data on the severity of conditions; this is a limitation common to most studies that employ data from routine health records. Routine health data also has the potential for bias due to differences in how healthcare staff record symptoms, diagnose conditions, or offer pharmacological therapies. If these are done less frequently for some groups then our analysis may have undercounted their actual number of conditions. This would lead to underestimation of the mortality risk associated with having more conditions. Our assessment of the contribution of new conditions emerging during follow-up was limited. We included the number of conditions at study start and end, which effectively models onset of new conditions as a covariate. Other studies have modelled transition to multimorbidity or to an additional condition separately from transition to death(42). Ethnic identity is related to several constructs such as migration status and preferred language, that may affect interaction with health care, ability to manage long-term conditions, and survival. These constructs are not well-captured in our data, but other studies have shown stark differences in mortality for people from minoritised ethnic groups that were born in the UK or Republic of Ireland and those that were born elsewhere (24,25).

## Conclusion

Given the finding of poorer survival with multiple conditions among some minoritised ethnic groups, work is now needed to identify factors that contribute to these inequalities. Within the health care setting, there may be opportunities to target clinical and self-management support. Our analysis suggests that attention to the management of circulatory and endocrine conditions may be particularly important for improving survival rates for minoritised ethnic groups with multiple conditions. Electronic health records can be further exploited to understand patterns of health care for these conditions. For example, analysis of the Myocardial Ischaemia National Audit Project found the multimorbidity cluster with the poorest survival was less likely to be receiving pharmacological therapies(43). Accounts from people with lived experience of multiple conditions also provide insight on the role of health professionals and health services to help people manage their conditions(37). This will include helping them to have a better understanding of their long-term conditions and the treatments they are recommended to adhere to, providing context-specific advice for lifestyle changes, and improving the way local services work together so that the health system is more straightforward to navigate.

In the longer term, solutions that prevent and delay the onset of multiple long-term conditions in communities that are ethnically or socioeconomically disadvantaged are needed. These will include local and national strategies that address the wider determinants of health within and beyond the health care system.

## Data Availability

The study uses data from the Clinical Practice Research Datalink (CPRD). CPRD data may be accessed by bona fide researchers for public health research which is funded by trustworthy organisations. New requests to access the data are assessed through a Research Data Governance process, https://www.cprd.com/Data-access
The full specification for the sample of data used for the current study is available from the authors upon request. The analytic code is available at https://github.com/HFAnalyticsLab/MLTCs_and_mortality_in_ethnic_groups

https://cprd.com/home

https://github.com/HFAnalyticsLab/MLTCs_and_mortality_in_ethnic_groups

## Acknowledgements

This work uses data provided by patients and collected by the NHS as part of their care and support. We also thank the Data Management Team at the Health Foundation.

