## Supplemental Tables 1 to 8 for "Associations between multiple long-term conditions and mortality in diverse ethnic groups"

**Supplementary Table 1. Long-term conditions counted in the current study**

| **Long-term condition** | **Body system** | **Time criteria (ever if not specified)** |  |
| --- | --- | --- | --- |
| Cancer | Cancers | In last 5 years |  |
| Atrial fibrillation | Diseases of the Circulatory System |  |  |
| Heart disease | Diseases of the Circulatory System |  |  |
| Heart failure | Diseases of the Circulatory System |  |  |
| Hypertension | Diseases of the Circulatory System |  |  |
| Stroke | Diseases of the Circulatory System |  | stroke |
| Vascular disease | Diseases of the Circulatory System |  |  |
| Diverticulosis | Diseases of the Digestive System |  |  |
| IBS | Diseases of the Digestive System |  |  |
| Liver disease | Diseases of the Digestive System |  |  |
| Hearing loss | Diseases of the Ear |  |  |
| Blindness | Diseases of the Eye |  |  |
| Diabetes | Diseases of the Endocrine System |  |  |
| Thyroid disorders | Diseases of the Endocrine System |  |  |
| Kidney disease | Diseases of the Genitourinary System |  |  |
| Asthma | Diseases of the Respiratory System | In last 12 months |  |
| Bronchiectasis | Diseases of the Respiratory System |  |  |
| COPD | Diseases of the Respiratory System |  |  |
| Viral hepatitis | Infectious Diseases |  |  |
| Alcohol misuse | Mental Health Disorders |  |  |
| Anxiety/depression | Mental Health Disorders | In last 12 months |  |
| Anorexia/bulimia | Mental Health Disorders |  |  |
| Dementia | Mental Health Disorders |  |  |
| Learning disability | Mental Health Disorders |  |  |
| Schizophrenia | Mental Health Disorders |  |  |
| Substance misuse | Mental Health Disorders |  |  |
| Arthritis | Musculoskeletal conditions |  |  |
| Epilepsy | Neurological conditions |  | epilepsy |
| Migraine | Neurological conditions |  |  |
| Multiple sclerosis | Neurological conditions |  |  |
| Parkinsons disease | Neurological conditions |  |  |
| Psoriasis | Skin conditions |  |  |

**Supplementary Table 2. Characteristics of analytical sample and excluded patients**

|  | Analytical sample  (% of patients) | Patients excluded due to missing data (% of patients) | Difference between analytical and excluded samples |
| --- | --- | --- | --- |
|  | N = 532059 | N=67941 |  |
| Baseline age |  |  | p<0.001 |
| 18-29y | 15.7 | 23.2 |  |
| 30-39y | 17.2 | 15.9 |  |
| 40-49y | 17.7 | 19.6 |  |
| 50-59y | 17.2 | 19.8 |  |
| 60-69y | 13.9 | 12.3 |  |
| 70-79y | 10.5 | 5.9 |  |
| 80+y | 7.8 | 3.4 |  |
| Women | 51.9 | 35.6 | p<0.001 |
| Index of Multiple Deprivation decile |  |  | p<0.001 |
| 1 (least deprived) | 10.7 | 15.0 |  |
| 2 | 10.3 | 12.5 |  |
| 3 | 10.5 | 11.9 |  |
| 4 | 10.0 | 10.6 |  |
| 5 | 9.6 | 9.5 |  |
| 6 | 9.9 | 8.9 |  |
| 7 | 10.4 | 9.9 |  |
| 8 | 9.7 | 7.6 |  |
| 9 | 10.1 | 7.6 |  |
| 10 | 8.7 | 6.7 |  |
| Complex multimorbidity |  |  |  |
| present | 11.1 | 3.3 | p<0.001 |
| Baseline number of conditions; mean(SD) | 1.16 (1.49) | 0.53 (0.97) | p<0.001 |

**Supplementary Table 3. Linear association between number of long-term conditions and mortality**

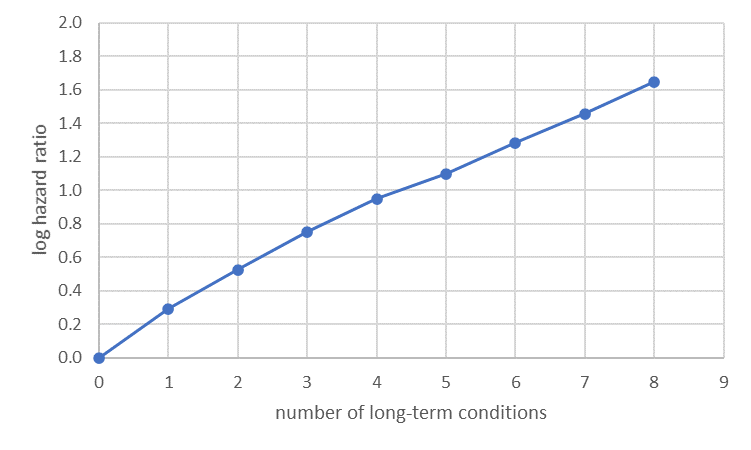

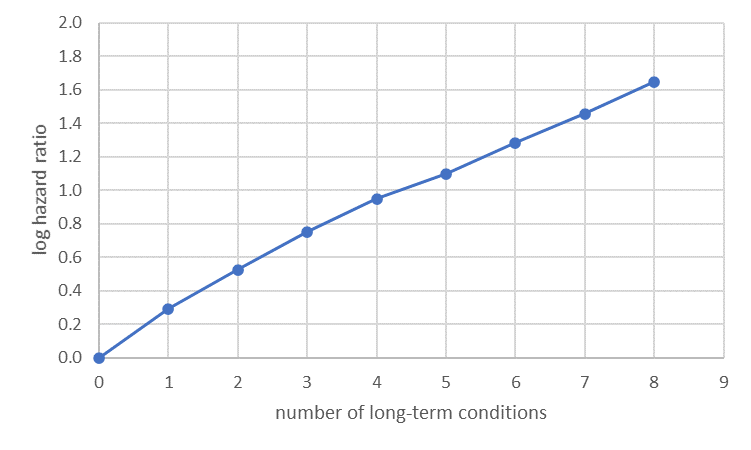

Figure shows log hazard ratio for death during follow-up by number of long-term conditions at baseline with 0 conditions as the reference group.

**Suppl Table 4. Characteristics of analytical sample by ethnicity**

|  | Bangladeshi | Pakistani | Indian | Other Asian | Chinese | Black African | Black Caribbean | Black Other | Mixed | Other | | White |
| --- | --- | --- | --- | --- | --- | --- | --- | --- | --- | --- | --- | --- |
| n | 3711 | 9233 | 15142 | 9022 | 4608 | 11517 | 7357 | 3119 | 7227 | 6910 | | 454213 |
| Baseline age N (%) | | | | | | | | | | | | |
| 18-29y | 1008 (27.2) | 2312 (25.0) | 2518 (16.6) | 1964 (21.8) | 1903 (41.3) | 2292 (19.9) | 1061 (14.4) | 849 (27.2) | 1988 (27.5) | 1496 (21.6) | | 66258 (14.6) |
| 30-39y | 1183 (31.9) | 2786 (30.2) | 4286 (28.3) | 2379 (26.4) | 1000 (21.7) | 2935 (25.5) | 1233 (16.8) | 780 (25.0) | 1905 (26.4) | 1928 (27.9) | | 71244 (15.7) |
| 40-49y | 826 (22.3) | 1976 (21.4) | 2943 (19.4) | 2131 (23.6) | 715 (15.5) | 3140 (27.3) | 1348 (18.3) | 632 (20.3) | 1474 (20.4) | 1628 (23.6) | | 77603 (17.1) |
| 50-59y | 312 (8.4) | 978 (10.6) | 2174 (14.4) | 1210 (13.4) | 446 (9.7) | 1985 (17.2) | 1783 (24.2) | 560 (18.0) | 1050 (14.5) | 975 (14.1) | | 80011 (17.6) |
| 60-69y | 199 (5.4) | 681 (7.4) | 1753 (11.6) | 799 (8.9) | 305 (6.6) | 673 (5.8) | 724 (9.8) | 137 (4.4) | 436 (6.0) | 475 (6.9) | | 67523 (14.9) |
| 70-79y | 123 (3.3) | 319 (3.5) | 937 (6.2) | 380 (4.2) | 140 (3.0) | 374 (3.2) | 661 (9.0) | 97 (3.1) | 240 (3.3) | 266 (3.8) | | 52338 (11.5) |
| 80+y | 60 (1.6) | 181 (2.0) | 531 (3.5) | 159 (1.8) | 99 (2.1) | 118 (1.0) | 547 (7.4) | 64 (2.1) | 134 (1.9) | 142 (2.1) | | 39236 (8.6) |
| Women N (%) | 1697 (45.7) | 4259 (46.1) | 7331 (48.4) | 4463 (49.5) | 2446 (53.1) | 5897 (51.2) | 3979 (54.1) | 1578 (50.6) | 3880 (53.7) | 3335 (48.3) | | 237282 (52.2) |
| Index of Multiple Deprivation N (%) | | | | | | | | | | |  | |
| 1 (least deprived) | 71 (1.9) | 291 (3.2) | 1130 (7.5) | 567 (6.3) | 335 (7.3) | 180 (1.6) | 112 (1.5) | 64 (2.1) | 449 (6.2) | 406 (5.9) | | 53518 (11.8) |
| 2 | 65 (1.8) | 299 (3.2) | 1126 (7.4) | 595 (6.6) | 379 (8.2) | 227 (2.0) | 139 (1.9) | 84 (2.7) | 421 (5.8) | 381 (5.5) | | 51305 (11.3) |
| 3 | 113 (3.0) | 399 (4.3) | 1174 (7.8) | 826 (9.2) | 422 (9.2) | 312 (2.7) | 168 (2.3) | 125 (4.0) | 500 (6.9) | 494 (7.1) | | 51265 (11.3) |
| 4 | 107 (2.9) | 448 (4.9) | 1234 (8.1) | 739 (8.2) | 395 (8.6) | 430 (3.7) | 270 (3.7) | 118 (3.8) | 463 (6.4) | 456 (6.6) | | 48316 (10.6) |
| 5 | 112 (3.0) | 594 (6.4) | 1451 (9.6) | 832 (9.2) | 336 (7.3) | 562 (4.9) | 410 (5.6) | 203 (6.5) | 604 (8.4) | 585 (8.5) | | 45523 (10.0) |
| 6 | 232 (6.3) | 863 (9.3) | 1907 (12.6) | 1048 (11.6) | 488 (10.6) | 943 (8.2) | 724 (9.8) | 259 (8.3) | 705 (9.8) | 596 (8.6) | | 44964 (9.9) |
| 7 | 321 (8.6) | 1128 (12.2) | 1922 (12.7) | 1286 (14.3) | 626 (13.6) | 1317 (11.4) | 980 (13.3) | 372 (11.9) | 862 (11.9) | 897 (13.0) | | 45448 (10.0) |
| 8 | 751 (20.2) | 1410 (15.3) | 2210 (14.6) | 1377 (15.3) | 667 (14.5) | 2268 (19.7) | 1325 (18.0) | 547 (17.5) | 1051 (14.5) | 1178 (17.0) | | 38932 (8.6) |
| 9 | 993 (26.8) | 1733 (18.8) | 1726 (11.4) | 1104 (12.2) | 573 (12.4) | 3188 (27.7) | 1753 (23.8) | 692 (22.2) | 1200 (16.6) | 1088 (15.7) | | 39877 (8.8) |
| 10 | 946 (25.5) | 2068 (22.4) | 1262 (8.3) | 648 (7.2) | 387 (8.4) | 2090 (18.1) | 1476 (20.1) | 655 (21.0) | 972 (13.4) | 829 (12.0) | | 35065 (7.7) |
| Complex multimorbidity present N (%) | 228 (6.1) | 679 (7.4) | 1233 (8.1) | 442 (4.9) | 117 (2.5) | 437 (3.8) | 848 (11.5) | 164 (5.3) | 326 (4.5) | 267 (3.9) | | 54332 (12.0) |
| Died during follow-up N (%) | 54 (1.5) | 184 (2.0) | 398 (2.6) | 132 (1.5) | 54 (1.2) | 135 (1.2) | 350 (4.8) | 46 (1.5) | 111 (1.5) | 104 (1.5) | | 29892 (6.6) |
| Follow-up time in years mean (SD) | 4.31 (1.41) | 4.31 (1.39) | 4.20 (1.48) | 4.03 (1.63) | 3.47 (1.90) | 4.05 (1.59) | 4.31 (1.36) | 4.16 (1.50) | 4.06 (1.59) | 4.00 (1.63) | | 4.25 (1.43) |
| Baseline number of long-term conditions (mean (SD) | 0.72 (1.17) | 0.82 (1.29) | 0.88 (1.32) | 0.63 (1.09) | 0.33 (0.81) | 0.57 (0.95) | 1.11 (1.42) | 0.63 (1.02) | 0.67 (1.05) | 0.54 (1.00) | | 1.23 (1.52) |
| Number of long-term conditions at end of follow-up (mean (SD) | 0.96 (1.39) | 1.07 (1.52) | 1.12 (1.55) | 0.83 (1.31) | 0.41 (0.97) | 0.74 (1.13) | 1.40 (1.67) | 0.83 (1.22) | 0.82 (1.24) | 0.71 (1.20) | | 1.55 (1.79) |

**Supplementary Table 5. Cox regression estimates from model 1 assuming no interaction between ethnicity and number of conditions**

| Covariate | Reference group | Regression estimate | Standard error | p-value |
| --- | --- | --- | --- | --- |
| Women | Men | -0.28 | 0.01 | p<0.001 |
| Baseline age | per 1 year increase, centred at age 50 | 0.10 | 0.00 | p<0.001 |
| Baseline number of conditions | per 1 condition increase | 0.29 | 0.01 | p<0.001 |
| Age x number of conditions | per 1 unit increase | -0.003 | 0.000 | p<0.001 |
| Ethnicity (main effect) | White (including white British, white Irish and other white) |  |  |  |
| Bangladeshi |  | -0.16 | 0.22 | p=0.4 |
| Pakistani |  | 0.08 | 0.13 | p=0.6 |
| Indian |  | -0.48 | 0.12 | p<0.001 |
| Other Asian |  | -0.30 | 0.16 | p=0.06 |
| Chinese |  | -0.90 | 0.32 | p=0.004 |
| Black African |  | -0.03 | 0.12 | p=0.8 |
| Black Caribbean |  | 0.19 | 0.13 | p=0.1 |
| Black other |  | 0.27 | 0.20 | p=0.2 |
| Mixed |  | 0.20 | 0.13 | p=0.1 |
| Other |  | -0.24 | 0.19 | p=0.2 |
| Age x ethnicity interaction: | per 1 year increase compared with White ethnicity |  |  |  |
| Bangladeshi |  | -0.004 | 0.009 | p=0.7 |
| Pakistani |  | -0.009 | 0.005 | p=0.09 |
| Indian |  | 0.007 | 0.004 | p=0.1 |
| Other Asian |  | -0.005 | 0.007 | p=0.5 |
| Chinese |  | 0.019 | 0.011 | p=0.07 |
| Black African |  | -0.015 | 0.007 | p=0.02 |
| Black Caribbean |  | -0.014 | 0.005 | p=0.002 |
| Black other |  | -0.030 | 0.010 | p=0.003 |
| Mixed |  | -0.024 | 0.006 | p<0.001 |
| Other |  | -0.002 | 0.008 | p=0.8 |

**Supplementary Table 6. Cox regression estimates from model 2 including interaction between ethnicity and baseline number of conditions and model 3 including possible explanatory factors**

|  | Model 2 |  |  | Model 3 |  |  |
| --- | --- | --- | --- | --- | --- | --- |
| Covariate | Regression estimate | Standard error | p-value | Regression estimate | Standard error | p-value |
| Women | -0.28 | 0.01 | p<0.001 | -0.29 | 0.01 | p<0.001 |
| Baseline age | 0.10 | 0.001 | p<0.001 | 0.10 | 0.001 | p<0.001 |
| Baseline number of conditions | 0.29 | 0.01 | p<0.001 | 0.28 | 0.01 | p<0.001 |
| Age x number of conditions | -0.003 | 0.0003 | p<0.001 | -0.003 | 0.0003 | p<0.001 |
| Ethnicity (main effect) |  |  |  |  |  |  |
| Bangladeshi | -0.10 | 0.22 | p=0.7 | -0.28 | 0.25 | p=0.2 |
| Pakistani | -0.26 | 0.15 | p=0.2 | -0.39 | 0.15 | p=0.01 |
| Indian | -0.54 | 0.12 | p<0.001 | -0.57 | 0.12 | p<0.001 |
| Other Asian | -0.41 | 0.16 | p=0.009 | -0.45 | 0.16 | p=0.003 |
| Chinese | -0.98 | 0.33 | p=0.002 | -0.98 | 0.33 | p=0.001 |
| Black African | -0.18 | 0.14 | p=0.2 | -0.34 | 0.14 | p=0.01 |
| Black Caribbean | 0.08 | 0.12 | p=0.5 | -0.04 | 0.12 | p=0.7 |
| Black other | -0.14 | 0.23 | p=0.5 | -0.28 | 0.24 | p=0.2 |
| Mixed | 0.11 | 0.16 | p=0.5 | 0.02 | 0.16 | p=0.9 |
| Other | -0.37 | 0.19 | p=0.06 | -0.45 | 0.19 | p=0.03 |
| Age x ethnicity interaction: |  |  |  |  |  |  |
| Bangladeshi | -0.002 | 0.010 | p=0.8 | -0.003 | 0.010 | p=0.8 |
| Pakistani | -0.019 | 0.006 | p=0.006 | -0.019 | 0.006 | p=0.002 |
| Indian | 0.005 | 0.004 | p=0.3 | 0.004 | 0.005 | p=0.4 |
| Other Asian | -0.010 | 0.007 | p=0.2 | -0.010 | 0.008 | p=0.2 |
| Chinese | 0.015 | 0.012 | p=0.2 | 0.014 | 0.011 | p=0.3 |
| Black African | -0.021 | 0.006 | p=0.003 | -0.021 | 0.006 | p=0.003 |
| Black Caribbean | -0.018 | 0.004 | p<0.001 | -0.019 | 0.004 | p<0.001 |
| Black other | -0.045 | 0.011 | p<0.001 | -0.044 | 0.011 | p<0.001 |
| Mixed | -0.027 | 0.007 | p<0.001 | -0.027 | 0.008 | p<0.001 |
| Other | -0.006 | 0.007 | p=0.4 | -0.006 | 0.008 | p=0.5 |
| Number of conditions x ethnicity interaction: |  |  |  |  |  |  |
| Bangladeshi | -0.03 | 0.07 | p=0.7 | -0.02 | 0.07 | p=0.8 |
| Pakistani | 0.15 | 0.04 | p<0.001 | 0.16 | 0.04 | p<0.001 |
| Indian | 0.04 | 0.03 | p=0.1 | 0.04 | 0.03 | p=0.1 |
| Other Asian | 0.08 | 0.05 | p=0.08 | 0.09 | 0.05 | p=0.06 |
| Chinese | 0.07 | 0.08 | p=0.4 | 0.07 | 0.08 | p=0.4 |
| Black African | 0.11 | 0.05 | p=0.03 | 0.11 | 0.05 | p=0.02 |
| Black Caribbean | 0.07 | 0.03 | p=0.01 | 0.07 | 0.03 | p=0.009 |
| Black other | 0.27 | 0.08 | p=0.006 | 0.27 | 0.08 | p=0.005 |
| Mixed | 0.07 | 0.08 | p=0.4 | 0.06 | 0.06 | p=0.4 |
| Other | 0.10 | 0.06 | p=0.05 | 0.10 | 0.06 | p=0.07 |
| Number of conditions at end of follow-up |  |  |  | 0.005 | 0.005 | p=0.5 |
| Index of Multiple Deprivation decile |  |  |  |  |  |  |
| 2 |  |  |  | 0.11 | 0.02 | p<0.001 |
| 3 |  |  |  | 0.15 | 0.02 | p<0.001 |
| 4 |  |  |  | 0.17 | 0.03 | p<0.001 |
| 5 |  |  |  | 0.17 | 0.03 | p<0.001 |
| 6 |  |  |  | 0.20 | 0.03 | p<0.001 |
| 7 |  |  |  | 0.29 | 0.03 | p<0.001 |
| 8 |  |  |  | 0.36 | 0.03 | p<0.001 |
| 9 |  |  |  | 0.44 | 0.03 | p<0.001 |
| 10 |  |  |  | 0.56 | 0.03 | p<0.001 |
| Likelihood ratio test | 21.8 on 10 df, 0.05>p>0.01 compared with model 1 | | | 392 on 10 df, p<0.001 compared with model 2 | | |

**Supplementary Table 7. Cox regression estimates from model including interaction between ethnicity and complex multimorbidity**

| Covariate | Regression estimate | Standard error | p-value |
| --- | --- | --- | --- |
| Women | -0.28 | 0.01 | p<0.001 |
| Baseline age | 0.10 | 0.001 | p<0.001 |
| Baseline complex multimorbidity | 0.85 | 0.03 | p<0.001 |
| Age x complex multimorbidity | -0.01 | 0.001 | p<0.001 |
| Ethnicity (main effect) |  |  |  |
| Bangladeshi | -0.18 | 0.21 | p=0.4 |
| Pakistani | -0.06 | 0.14 | p=0.7 |
| Indian | -0.48 | 0.11 | p<0.001 |
| Other Asian | -0.40 | 0.15 | p=0.01 |
| Chinese | -1.05 | 0.30 | p<0.001 |
| Black African | -0.20 | 0.13 | p=0.1 |
| Black Caribbean | 0.14 | 0.12 | p=0.3 |
| Black other | -0.11 | 0.22 | p=0.6 |
| Mixed | 0.16 | 0.13 | p=0.2 |
| Other | -0.37 | 0.19 | p=0.05 |
| Age x ethnicity interaction: |  |  |  |
| Bangladeshi | -0.003 | 0.009 | p=0.8 |
| Pakistani | -0.013 | 0.006 | p=0.02 |
| Indian | 0.007 | 0.004 | p=0.1 |
| Other Asian | -0.006 | 0.008 | p=0.5 |
| Chinese | 0.012 | 0.011 | p=0.3 |
| Black African | -0.018 | 0.007 | p=0.008 |
| Black Caribbean | -0.014 | 0.005 | p=0.003 |
| Black other | -0.045 | 0.011 | p<0.001 |
| Mixed | -0.020 | 0.007 | p=0.004 |
| Other | -0.002 | 0.008 | p=0.8 |
| Complex multimorbidity x ethnicity interaction: |  |  |  |
| Bangladeshi | 0.01 | 0.28 | p=0.9 |
| Pakistani | 0.45 | 0.16 | p=0.004 |
| Indian | 0.01 | 0.11 | p=0.9 |
| Other Asian | 0.23 | 0.21 | p=0.3 |
| Chinese | 0.62 | 0.30 | p=0.04 |
| Black African | 0.46 | 0.17 | p=0.009 |
| Black Caribbean | 0.06 | 0.10 | p=0.6 |
| Black other | 1.15 | 0.33 | p<0.001 |
| Mixed | -0.22 | 0.25 | p=0.4 |
| Other | 0.21 | 0.21 | p=0.3 |

**Supplementary Table 8. Cox regression estimates from model including interaction between circulatory/endocrine/respiratory conditions and mortality**

|  | Circulatory condition | | | Endocrine condition | | | Respiratory condition | | |
| --- | --- | --- | --- | --- | --- | --- | --- | --- | --- |
| Covariate | Regression estimate | Standard error | p-value | Regression estimate | Standard error | p-value | Regression estimate | Standard error | p-value |
| Women | -0.27 | 0.01 | p<0.001 | -0.29 | 0.01 | p<0.001 | -0.29 | 0.01 | p<0.001 |
| Baseline age | 0.10 | 0.0007 | p<0.001 | 0.10 | 0.0006 | p<0.001 | 0.11 | 0.0005 | p<0.001 |
| Baseline condition present | 0.12 | 0.03 | p<0.001 | 0.43 | 0.03 | p<0.001 | 0.71 | 0.03 | p<0.001 |
| Age x condition present | 0.008 | 0.001 | p<0.001 | -0.005 | 0.001 | p<0.001 | -0.011 | 0.001 | p<0.001 |
| Ethnicity (main effect) |  |  |  |  |  |  |  |  |  |
| Bangladeshi | -0.26 | 0.22 | p=0.2 | -0.38 | 0.22 | p=0.1 | -0.03 | 0.19 | p=0.9 |
| Pakistani | -0.14 | 0.14 | p=0.3 | -0.25 | 0.16 | p=0.1 | 0.11 | 0.14 | p=0.4 |
| Indian | -0.53 | 0.12 | p<0.001 | -0.63 | 0.12 | p<0.001 | -0.43 | 0.11 | p<0.001 |
| Other Asian | -0.54 | 0.16 | p=0.001 | -0.46 | 0.16 | p=0.005 | -0.32 | 0.16 | p=0.04 |
| Chinese | -1.13 | 0.31 | p<0.001 | -1.14 | 0.32 | p<0.001 | -1.01 | 0.31 | p=0.001 |
| Black African | -0.26 | 0.13 | p=0.05 | -0.32 | 0.13 | p=0.02 | -0.01 | 0.12 | p=0.9 |
| Black Caribbean | 0.05 | 0.13 | p=0.7 | 0.08 | 0.13 | p=0.5 | 0.23 | 0.12 | p=0.07 |
| Black other | -0.19 | 0.24 | p=0.4 | -0.13 | 0.23 | p=0.6 | 0.13 | 0.21 | p=0.5 |
| Mixed | 0.04 | 0.14 | p=0.8 | 0.06 | 0.13 | p=0.6 | 0.21 | 0.14 | p=0.1 |
| Other | -0.50 | 0.19 | p=0.007 | -0.36 | 0.19 | p=0.06 | -0.25 | 0.19 | p=0.2 |
| Age x ethnicity interaction: |  |  |  |  |  |  |  |  |  |
| Bangladeshi | -0.003 | 0.010 | p=0.8 | -0.007 | 0.010 | p=0.5 | -0.004 | 0.008 | p=0.6 |
| Pakistani | -0.014 | 0.006 | p=0.01 | -0.014 | 0.006 | p=0.02 | -0.010 | 0.005 | p=0.05 |
| Indian | 0.007 | 0.004 | p=0.1 | 0.007 | 0.004 | p=0.1 | 0.006 | 0.004 | p=0.2 |
| Other Asian | -0.006 | 0.008 | p=0.4 | -0.002 | 0.007 | p=0.7 | -0.003 | 0.007 | p=0.6 |
| Chinese | 0.017 | 0.012 | p=0.2 | 0.015 | 0.011 | p=0.2 | 0.019 | 0.010 | p=0.07 |
| Black African | -0.013 | 0.007 | p=0.06 | -0.019 | 0.007 | p=0.005 | -0.015 | 0.006 | p=0.02 |
| Black Caribbean | -0.014 | 0.005 | p=0.004 | -0.015 | 0.005 | p=0.003 | -0.013 | 0.004 | p=0.004 |
| Black other | -0.041 | 0.013 | p=0.002 | -0.040 | 0.011 | p=0.002 | -0.027 | 0.010 | p=0.007 |
| Mixed | -0.020 | 0.007 | p=0.006 | -0.024 | 0.007 | p<0.001 | -0.022 | 0.006 | p<0.001 |
| Other | -0.006 | 0.008 | p=0.5 | -0.003 | 0.008 | p=0.9 | -0.002 | 0.007 | p=0.8 |
| Condition present x ethnicity interaction: |  |  |  |  |  |  |  |  |  |
| Bangladeshi | 0.23 | 0.34 | p=0.5 | 0.40 | 0.30 | p=0.2 | -0.28 | 0.31 | p=0.4 |
| Pakistani | 0.56 | 0.18 | p=0.002 | 0.67 | 0.15 | p<0.001 | 0.11 | 0.18 | p=0.5 |
| Indian | 0.10 | 0.12 | p=0.4 | 0.22 | 0.11 | p=0.04 | 0.04 | 0.12 | p=0.8 |
| Other Asian | 0.36 | 0.22 | p=0.1 | 0.10 | 0.19 | p=0.6 | -0.12 | 0.22 | p=0.6 |
| Chinese | 0.20 | 0.38 | p=0.6 | 0.46 | 0.28 | p=0.1 | 0.10 | 0.42 | p=0.8 |
| Black African | 0.15 | 0.20 | p=0.5 | 0.51 | 0.18 | p=0.005 | -0.80 | 0.34 | p=0.02 |
| Black Caribbean | 0.16 | 0.15 | p=0.3 | 0.14 | 0.13 | p=0.3 | -0.30 | 0.12 | p=0.01 |
| Black other | 0.84 | 0.41 | p=0.04 | 0.96 | 0.31 | p=0.002 | 0.27 | 0.36 | p=0.4 |
| Mixed | 0.06 | 0.24 | p=0.8 | 0.16 | 0.24 | p=0.5 | -0.43 | 0.26 | p=0.09 |
| Other | 0.42 | 0.23 | p=0.06 | -0.02 | 0.21 | p=0.9 | -0.40 | 0.29 | p=0.2 |
